# Perspectives on the enhancement of commercially available antibiotics by natural products

**DOI:** 10.1101/2022.08.22.22279086

**Authors:** Lucia Nitsch-Velasquez, Sara B. Barrios, Ricardo A. Montoya, Rosy Canales

## Abstract

**Background:** Opportunistic resistant bacteria are health and economically relevant in the health care systems and in industries worldwide, especially in the so-called resistant bacteria era (RBE). Enhancing the activity of commercially available antibiotics (CAAs) with different types of natural products (NPs) is a successful antimicrobial strategy, for instance the amoxicillin and clavulanate mixture.

**Objective:** To find research trends in this field during 2015-2020 and to detect potential drug hits with potential to diversify formulations and materials design that can be useful to manage the RBE.

**Systematic review results:** It yielded 190 reports of synergistic effects of CAAs and NPs. The analyzed variables were: a) natural products origin: plant family, genera, secondary metabolite type; b) strains: +/- Gram, genera, most frequent species, application field; and c) CAAs: family, most frequent CAAs. The families with potential to have more bioactive species were *Apocynaceae, Rubiaceae, Euphorbiaceae* (I_*sbio*_ factor). *Lonicera* had the highest reports amount.

Polyphenols and flavonoids were the majority of pure NPs tested. Several potential drug hits for antibiotic activity enhancement at synergistic level were identified together with potential mechanisms of action: berberine (drug efflux inhibitor–DEI, biofilm inhibitor–BI), curcumin (BI), essential oils (BI), 3-o-metyl-butylgallato (inhibition of fatty acid saturation), among others. About the half of the tested strains were gram positive, being Methicillin Resistant *Staphylococcus aureus* (MRSA) the most frequently tested. *Escherichia coli* was the gram negative strain most frequently reported, including enterotoxigenic and extended spectrum beta-lactamases producers. The growth of other foodborne genera strains, such as *Listeria* and *Salmonella*, were also inhibited. Aminoglycosides were the family most reported, with gentamicin as the most commonly studied.

**Conclusions:** NPs as either as plant extracts from a variety of families, or as purified compounds specially flavonoids and polyphenols, have shown effective results to enhance the antibiotic activity of CAAs against gram positive and negative strains relevant to HC and FI. Their mechanisms of action are starting to be determined, as the case EPIs and BIs. Further research is needed to achieve co-formulations and materials design useful for those fields, that can certainly be positively impacted by pursuing this strategy.

## Background

Opportunistic bacteria are relevant in the health care systems and in industries that involve living organisms. For instance, in 2021 the cost of human illness caused by food borne pathogens costed more than 15.6billion USD to USA. A 48% of those outbreaks were related to meat, and 34% were related to plant based foods. This phenomena contributes to the emergence of antibiotic-resistant bacteria which positively feed backs the increment of food borne antibiotic-resistant infections. Among the CDC and FDA, and USDA strategies to address these issues are: **a)** to stimulate the antibiotic drug discovery, **b)** to improve the appropriate antibiotic use in veterinary medicine and agriculture, and **c)** to ensure that the related industries have tools, information, and training on antibiotic use [1, 2, 3, 4, 5].

To properly manage the resistant bacteria era, humankind must advance its antimicrobial toolbox. One strategy is to enhance the activity of already commercialized antibiotics, and even to revert antibacterial resistant by co-formulating the antibiotic drugs with those enhancers. A successful example of this approach is the mixture of amoxicillin and clavulanic acid. This mixture was patented in 1985, consisting of a semi-synthetic derivative of penicillin mixed with an inhibitor of the enzyme beta-lactamase isolated from *Streptomyces clavuligerus*. The market for amoxicillin is expected to raise up to 4,256 millioun USD by 2026, at the same time resistant strains are emerging and antibiotic drug discovery and re-formulation is on demand (CDC) [6, 3].

Acknowledging the success of amoxicillin-clavulanate potassium combo and the relevance of the resistant bacteria in health care systems and industry, and taking in account the promise of natural products as source of bioactive compounds, a systematic literature review was performed aiming to identify potential natural products that can improve the antibiotic activity against opportunistic bacteria, with potential applications in the food industry.

## Literature Review Methods

### Literature search parameters

The literature search parameters were defined as following: **i) Databases and search engines:** National Center for Biotechnology Information (NCBI) – Pubmed Central,[7] Scifinder,[8] In the cases in which the search engine also yielded recommended articles related to the found article, follow up of such studies was performed.

**ii) Publication date**: in the range from april 2016 up to 2020 **iii) Targeted content**: Antimicrobial activity evaluation of commercially available antibiotics together with natural extracts against opportunistic microbes (CAAs and NE–OM), such that yielded synergistic effects results which data analysis included either FICI or a statistical comparison between control and test groups. Also, analog results from testing the main component(s) of any given natural extract were also included.

**iv) Keywords and phrases**: The keywords applied to start the literature search in the different databases and search engines were: synergistic effects natural products and antibiotics, botanicals and antibiotics bioassays, plant extracts interaction with antibiotics, and antibiotic adjuvant bioassays. **v) Exclusion criteria**: results classified as either antagonist, additive, or non–interaction effects of CADs and NPs tests; results classified as either enhancement or modulating effects of CADs and

NPs tests, such that were reported without statistical analysis, such that it was not possible to conclude if synergistic effects were observed; only the NPs (either as extract or purified components from it) were tested for antimicrobial activity; mixture of two or more NPs, even if those yielded antimicrobial synergistic effects; mixture of two or more CADs, even if those yielded antimicrobial synergistic effects.

### Data analysis

The selected registries were analyzed based on a set of parameters that are analyzed in the following paragraphs. In brief, first, the analysis of the origin was divided into plant extracts and type of secondary metabolite. To further analyze the extracts origin, the frequency of reported families and genus were tabulated. In order to detect potential mining taxa for either more bioactive species (**I**_*sbio*_ above 0.80 for a given family) or to define species that are the most studied (**I**_*sbio*_below 0.3 for a given family), we proposed and utilized the **I**_*sbio*_ index for further analysis (see Fig. 1).

**Figure 1.**
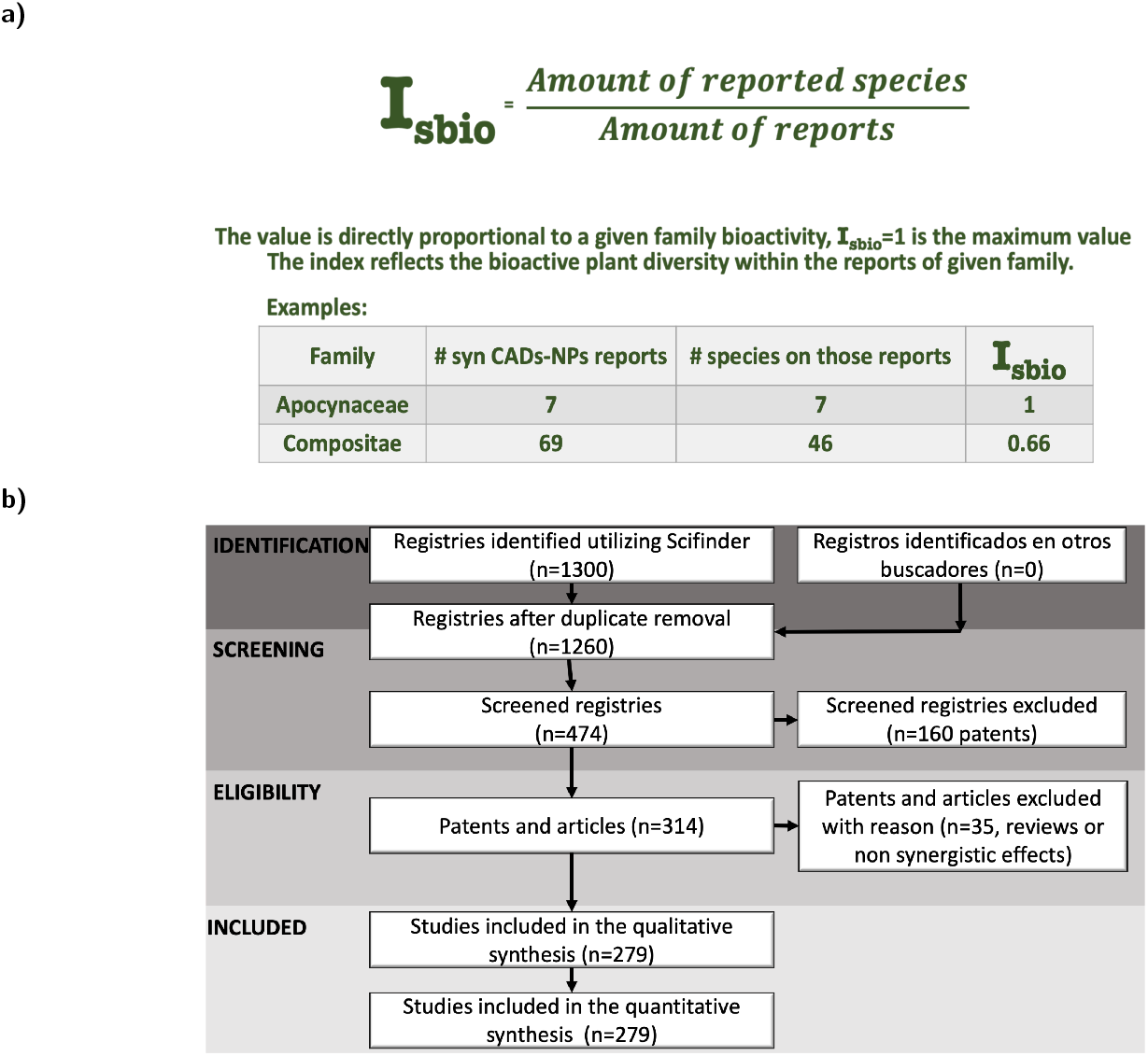
**a)** Definition of **I**_*sbio*_]Definition of **I**_*sbio*_. **b)** Prisma scheme of the systematic literature review of antibiotic synergistic assays of NPs and CAAs. Results of the search are availbale upon request.

Then, the tested bacteria were analyzed by strain, gram positive or negative, and field of relevance. The antibiotics that rendered synergistic effects were grouped by their type. Additionally, the natural products that yielded antibiofilm inhibition activity were compiled.

## Results and Discussion

A succinct summary is shown in Fig. 3 and the prisma escheme for the literature review in Fig. 1. A total of 270 reports of synergistic effects of natural products and commercially available antibiotics (Syn-NPs-CAAs) were retrieved, together with several reviews on the topic [9, 10, 11, 12, 13, 14, 15, 16, 17, 18, 19, 20, 21, 22, 23, 24, 25].

The Syn-NPs-CAAs were referred as a combinatory therapy [26], Chinese medicine and Western medicine integration [27], and an hybrid combination [15], all of them highlighting the fact of the utilization of an already validated commercially available antibiotic and a natural product that has not yet been validated by the same means, but that is known to be active in traditional medicine. Such a natural product can range from a pure compound, to fractionated extracts of a given species, to a mixture of extracts from several species.

The Syn-NPs-CAAs approach may be useful in the food industry for design of new packaging or for switching bacteria control to this hybrid formulation[15]. The synergistic effects were observed in studies with slightly variants regarding study focus, these are commented accordingly in the text.

The selected registries were analyzed based on a set of parameters that are analyzed in the following paragraphs. In brief, first, the analysis of the origin and type of natural product is presented in Sec., also in Figs. 4, 4, and 2, and Table 4. Then, the tested bacteria were analyzed by gram positive or negative, strain, and field of relevance, and field, see Sec. and 5. The antibiotics that rendered synergistic effects were grouped by their type are shown in Fig. 5. Additionally, the natural products that yielded antibiofilm inhibition activity (Sec.) were compiled in Figs. 2 and 1. The chapter closes with remarks and future prospects for research, Sec., development and innovation utilizing natural products as enhancers of antibiotic activity of CAAs.

**Figure 2.**
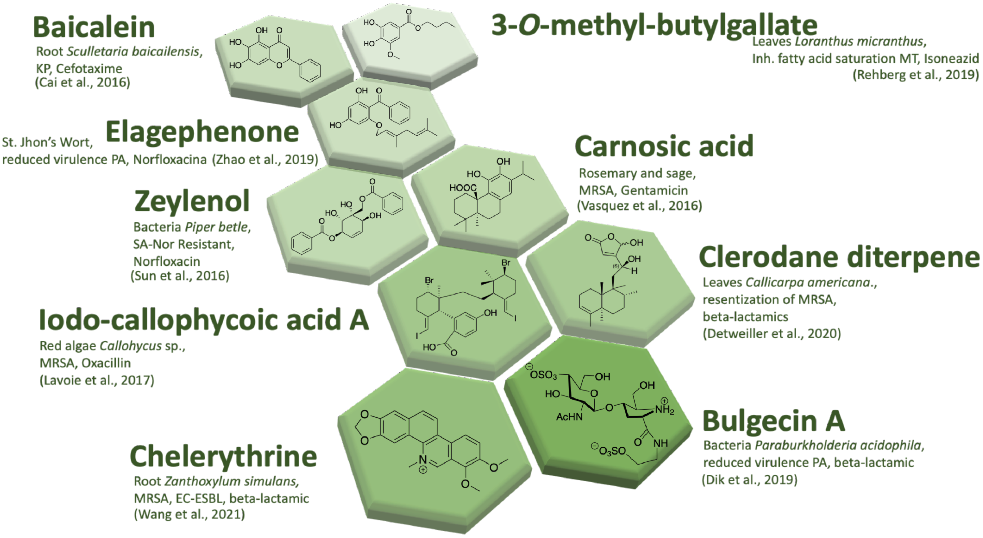
Examples of metabolites that yielded synergistic effects with CAAs and opportunistic bacteria.

**Figure 3.**
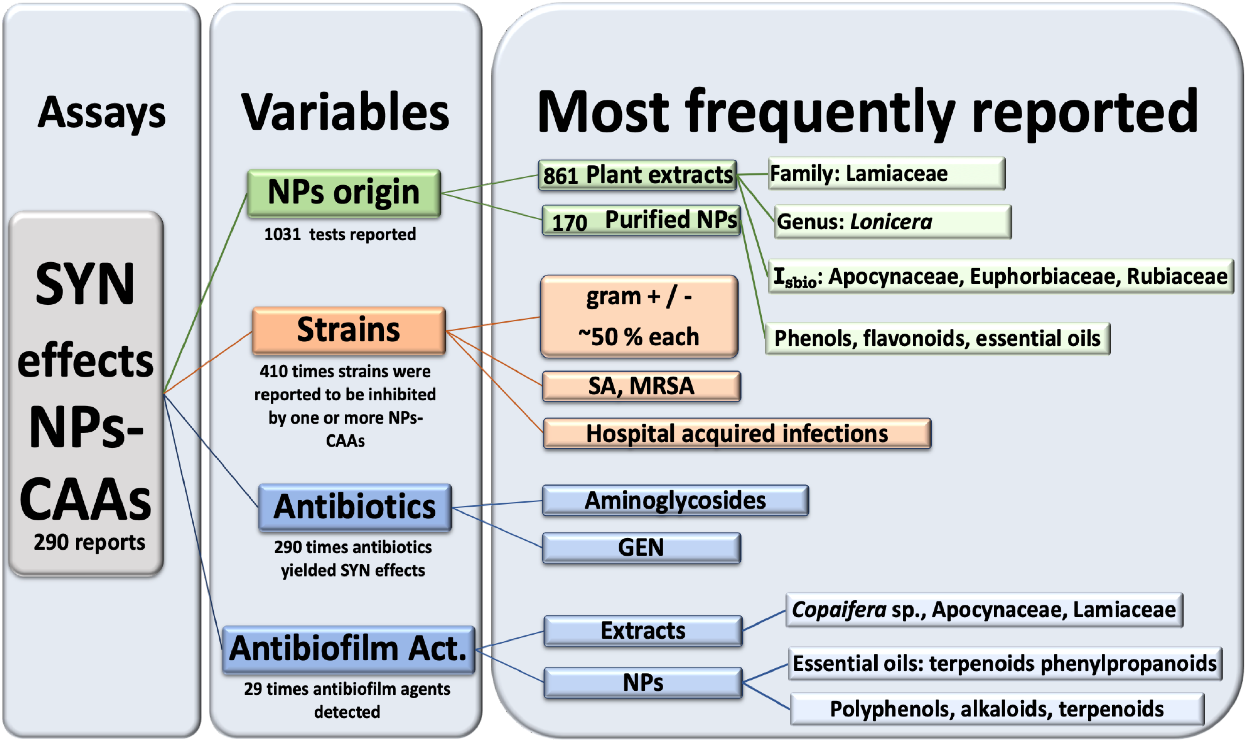
Summary of results of 279 reports of antibiotic synergistic assays of NPs and CAAs.

**Figure 4.**
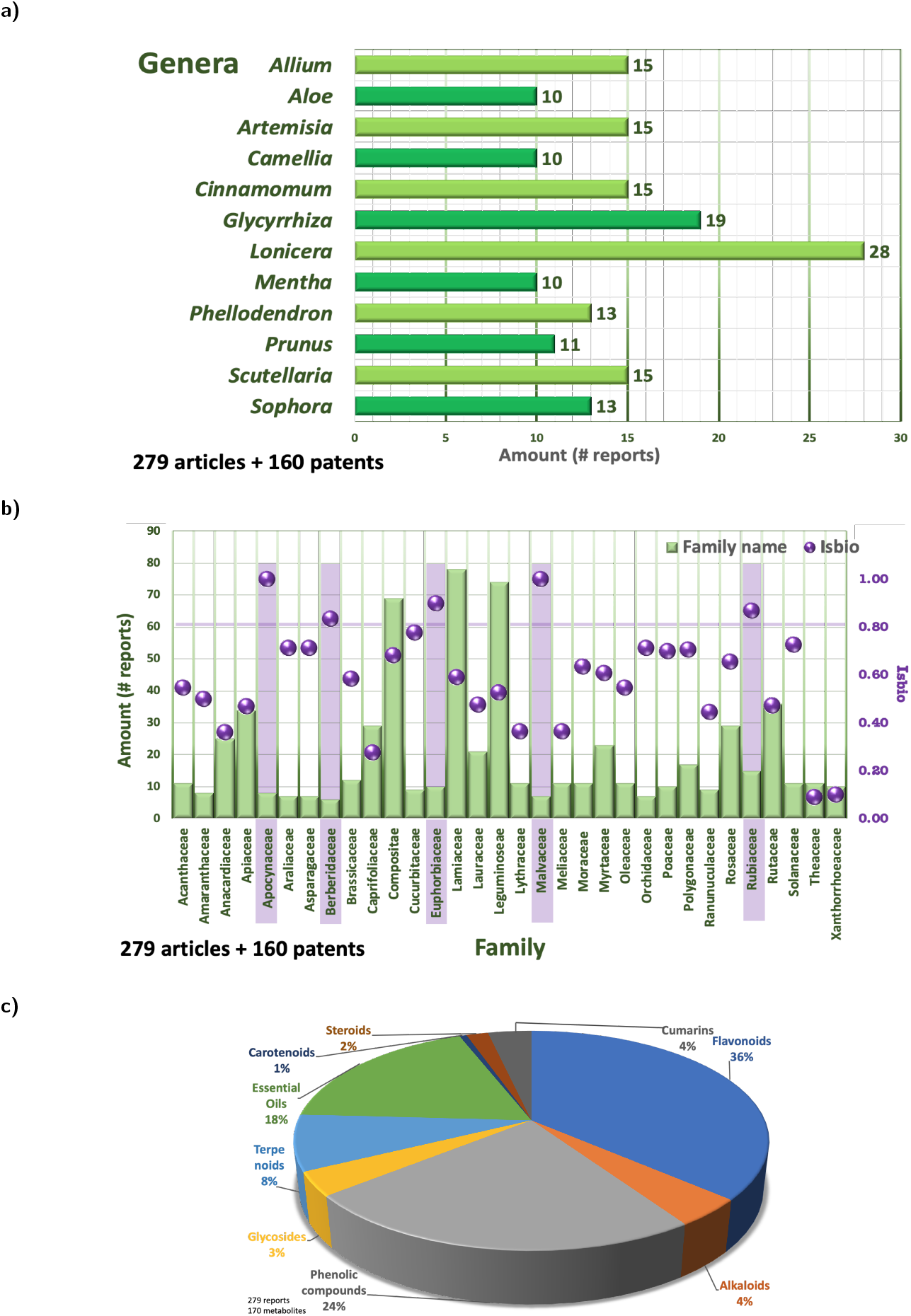
Distribution of the plant genera and families in 439 retrieved registries of antibiotic synergistic assays of NPs and CAAs, including reports and patents. **a)** Distribution of the plant genera. **b)**Distribution of the plant families with more than six reports and their **I**_*sbio*_, index range of 0.80 and 1 is highlighted. **c)** Secondary metabolite distribution of the 170 pure compounds tested

**Figure 5.**
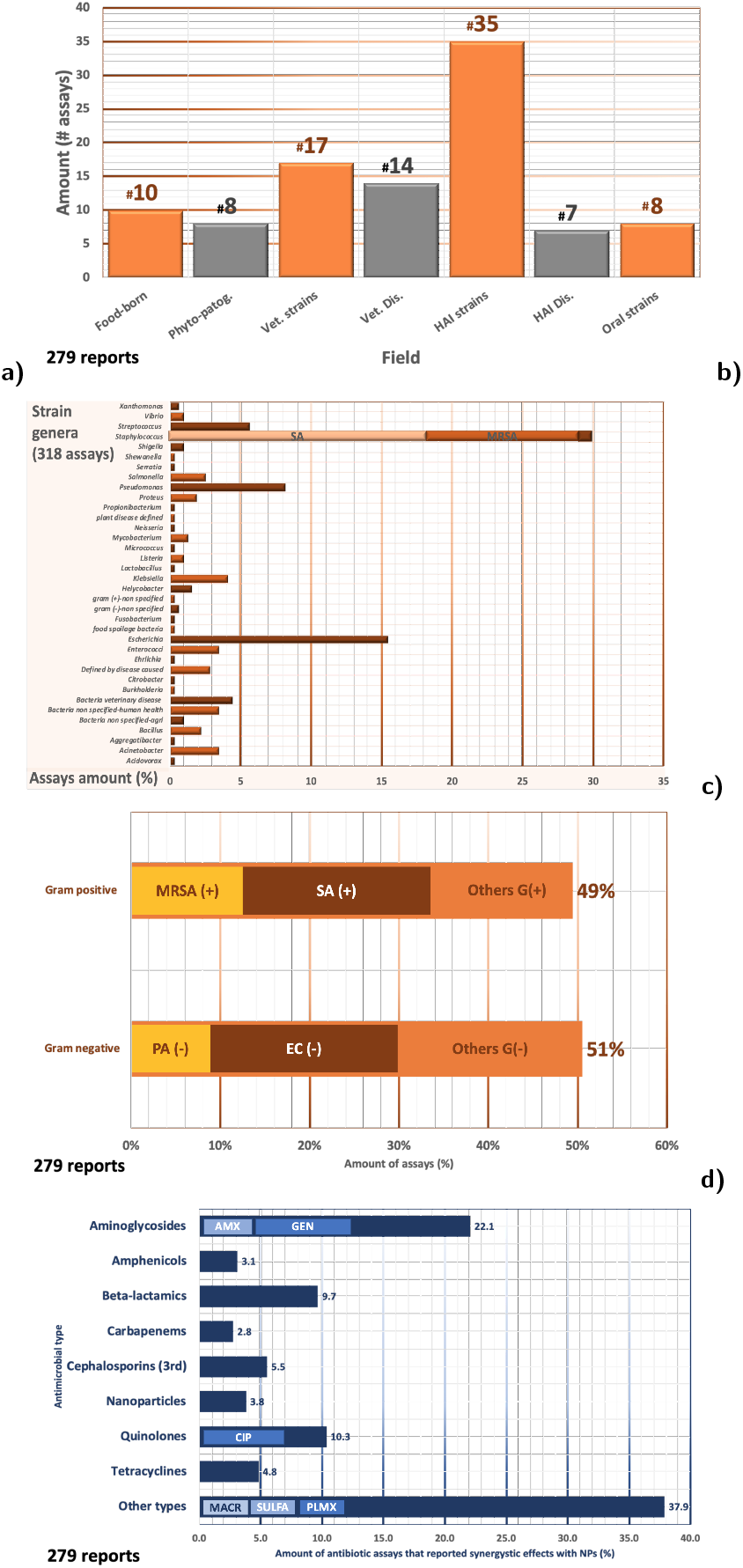
Distribution of strains and commercially available antibiotics. **a)** Distribution of the 99 different strains reported grouped by field in 279 reports of antibiotic synergistic assays of NPs and CAAs. **b)** Distribution of tested strains by their Gram dying. **c)** Distribution of tested strains grouped by genera. **d)** Distribution of the commercially available antibiotics utilized across those studies.

### Origin and type of the natural products

The species studied covered from edible plants, those used in traditional medicine, up to weeds. The genera *Lonicera* (Capriofoliaceae) was reported more frequently, being almost the only genera explored in that family, notice the **I**_*sbio*_ of 0.28, see Fig. 4 and 4. Green tea *Camelia sinensis* (Theaceae) and *Aloe vera* (Xanthorrhoeaceae) are the most studied species of their respective families, and it seems as no other species are currently under the radar of synergistic effects research (**I**_*sbio*_ below 0.20).

According to an **I**_*sbio*_ above 0.80, see Fig. 4, among the families of interest due to their potential to find bioactive species are Apocynaceae, Euphorbiaceae, Malvaceae and Rubiaceae [22]. The Compositae family is also of interest to explore more species, even though its current **I**_*sbio*_ is 0.68. Compositae is one of the largest among the plant families, actually is the major group of flowering plants, with more than 27,000 known species [28].

A total of 290 assays of secondary metabolites yielded synergistic interactions with CAAs against opportunistic bacteria, some examples are given in 4 and 1. Their distribution by secondary metabolite type is shown in Fig. 2. Flavonoids and phenolics represented the majority of the tested purified extracts. The flavonoids and polyphenols families may play an important role increasing the antibiotic bioavailability, and might become relevant in hybrid formulations and materials design, *e*.*g*., with CAAs [29, 30, 31, 32, 33].

Terpenoids as merulinic acid and a ursolic acid glycoside, among others, damaged the bacterial cell wall [34, 35]. Berberine can be considered a drug lead for efflux pump inhibition, such as berberine, zeylenol and bulgecin A, see Table 4 and references therein. Currently, berberine main limitation is its low bioavailability in the body [36, 37, 38, 39, 40, 32, 41, 42].

Other examples of efflux pump inhibitors are sophoroflavone G, jatrorrhizine, isovaleryl shikonin, griseviridin, 2-(2-aminophenyl)indole, flavonoids, essential oils and several plant extracts. It should be mentioned that both berberine and jatrorrhizine have also been isolated from *Mahonia bealei*, together with a variety of other alkaloids, terpenoids and polyphenols, and synergistic interactions with CAAs can be an expected result [43, 44, 45, 46, 47, 48, 40, 49, 50, 51, 52, 53].

### Strains

The mixture of NPs and CAAs were effective against strains which distribution is presented in Fig. 5. Opportunistic bacterial strains were the center of the Fractional Inhibitory Concentration Index—FICI assays. FICI was determined applying the checkerboard method (except for at least five studies that compared statistical difference by *p* value). Among the bioactivities assayed were minimal inhibitory concentration, time kill assay, and biofilm inhibitory concentration. The origin of the strains included ATCC with a variety of resistant genes and other commercially available sources, as well as clinical isolates. First line antibiotics were commonly explored, see Fig. 5, with aminoglycosides being majority.

The most frequently reported genera was *Staphylococcus*, mostly *S. aureus* (SA) and MRSA. The other two main strains were the gram negative *Escherichia coli* (EC), including ESBL-producing EC and enterotoxigenic EC, and *Pseudomonas aeruginosa* (PA) including MDR variants. MRSA is in the WHO list of High Priority Bacteria that requires research and development of new antibiotics[54], and the CDC classifies MRSA as Serious Threat Level [55]. In 2014, MRSA was included among the *US government National Targets for Combating Antibiotic-Resistant Bacteria*, aiming to reduce by half (to—at least—50%) the bloodstream infections caused by MRSA [56]. Several of the studied strains belong to the so–called ES-KAPE pathogens species (**E***nterococcus faecium*, **S***taphylococcus aureus*, **K***lebsiella pneumoniae*, **A***cinetobacter baumanii*, **P***seudomonas aeruginosa*, **E***nterobacter*), a set of antibiotic–resistant pathogenic bacteria that represents new paradigms regarding pathogenesis, transmission, and resistance [57].

A set of 35 studies were focused in the type of infections more than its in the causal agent, for instance foodborne and oral infections, and those related to chronic inflammatory diseases, and veterinary, especially poultry and livestock [58, 59, 60, 61, 62, 63, 64, 65, 66, 67, 68, 69, 70, 71, 72, 73, 74, 75, 76, 77, 78, 79, 80, 81, 82, 83, 84].

The growth of foodborne strains *Listeria* sp., *Salmonella* sp., *Vibrio* sp., *Shigella* sp. was inhibited by a variety of NPs-CAAs, including essential oils such as thymol and nerolidol [85, 86, 87, 88, 89, 90, 91, 92, 93, 94, 31, 95, 96, 27, 97, 98, 48, 40, 99, 100, 101, 102, 103, 104, 105, 106, 107, 70, 108, 86].

Oral infections caused by opportunistic bacteria are an active target for drug discovery, including the virulent factors modulation such as the biofilm formation (see below Sec.) and a set of mouthwashes and toothpastes formulations had been patented [25, 53, 109, 110, 111, 112, 113, 114, 115, 24].

Another target, at bioassay level, was the gut microbiota regulation through the intake of selected probiotics [116]. Inhibition of the growth of gram negative bacteria represented the half of the reported strains, including MDR variants. Examples of natural products inhibiting EC, KP, and PA can be found in Fig. 4. Edible properties are relevant, especially for safety concerns on utilizing plant extracts. An example of the 49 Syn-NPs-CAAs for EC is the methanolic extracts of edible plants *Psidium guajava* L., *Persea americana* Mill., *Camellia sinensis* L., *Mangifera indica* L., *Coula edulis* Baill., and *Citrus sinensis* L. [93]. Other plant extracts inhibited EC growth with focus on foodborne, hospital acquired infections and veterinary [117, 86, 29, 118, 119, 120, 30, 121, 122, 27, 123, 124, 100, 125, 126, 127, 128, 129, 92, 130, 48, 131, 132, 93, 133, 134, 135, 136, 137, 138, 99, 139, 93, 140, 141, 142, 143, 144, 95, 89, 145, 146, 147, 78, 80, 81, 82, 93].

### Inhibition of Toxin Production and Antibiofilm Activities

Among the reported mechanisms of action that renders synergistic effects of NPs and CAAs are the interaction of NPs with bacteria virulent factors. Which included inhibition of toxin production, biofilm formation, interference with quorum sensing molecules, inhibition of penicillin binding proteins, pump efflux inhibitors and pore forming compounds [148, 149, 150, 151, 49, 152, 50, 34, 35].

Essential oils and flavonoids are among the set of drug hits for inhibiting enterotoxin production *in vitro*. Certain polyphenols inhibited the production of enterotoxins by *S. aureus* MDR [103]. And 5-Hydroxy-3,7,4’-trimethoxyflavone inhibited the enterotoxin production by *E. coli* [141, 120]. The extract of *Spondias mombin* L. (Anacardiaceae) leaves also enhanced the amoxicillin effect against enterotoxic EC strains, and further research may also lead other drug hits for this activity [141]. Further research on these natural products may lead to improve therapies to treat hemorrhagic diarrhea infections that are of high relevance in lower income countries [2].

Biofilm has been identified as a critical point for foodborne bacteria, as they are related to harbor variants that are resistant to antibiotics and cleaning products [4]. Antibiofilm activity was specifically reported, and usually detected with ethidium bromide assay or electron microscopy [50, 153]. The plant extracts that inhibited biofilm formation are listed in Table 2. For instance, the essential oils from the weed *Mikania cordifolia*, especially limonene, enhanced the CAAs activity against foodborne bacteria, probably via biofilm formation inhibition [89]. Specific polyphenols, alkaloids and terpenoids have also inhibited biofilm formation, see Table 1. They can be envisioned as drug hits for antibiofilm activity [154, 155, 36, 156, 157, 158, 159, 160, 161].

**Table 1.**
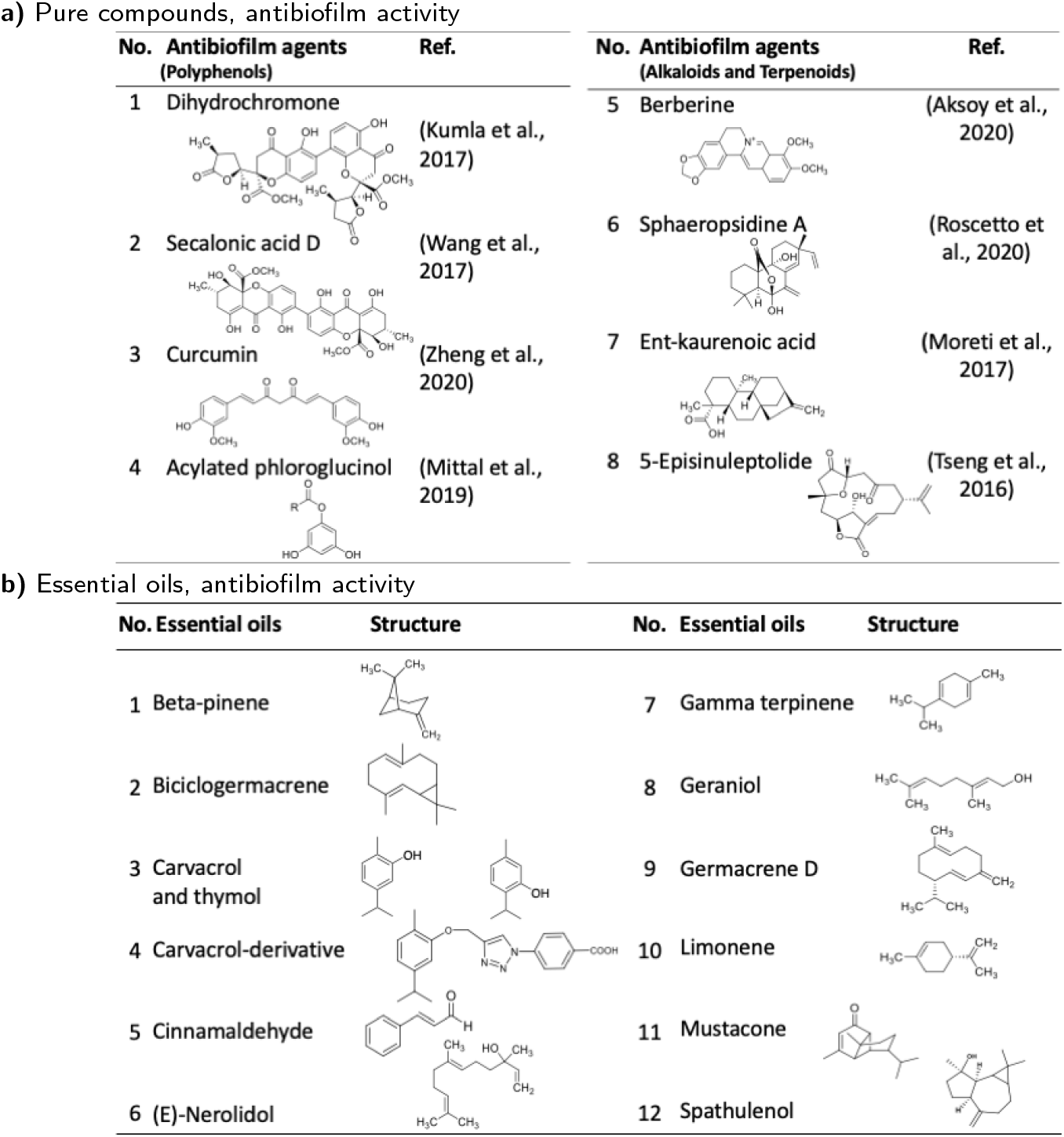
Antibiofilm agents. **a)** Polyphenols, alkaloids and terpenoids, as pure compounds, that yielded antibiofilm activity. **b)** Essential oils and an EO’s derivative, that yielded antibiofilm activity.

**Table 2.** Species of the plant extracts that yielded antibiofilm activity.

| No. | Inhibition of biofilm formation by extracts from | Family | Isbio | Refs. |
| --- | --- | --- | --- | --- |
| 1 | *Ajuga bracteosa* (silver nanoparticles) | Lamiaceae | 1.7 | (Nazer et al., 2020) |
| 2 | *Azadirachta indica* | Meliaceae | 2.8 | (Quelemes et al., 2015) |
| 3 | *Cinnamomum tamala* | Lauraceae | 2.1 | (Rubini et al., 2018) |
| 4 | *Copaifera duckei* | Leguminosae | 1.9 | (Goncalvez et al., 2018) |
| 5 | *Copaifera pubiflora* | Leguminosae | 1.9 | (da Silva et al., 2020) |
| 6 | *Copaifera trapezifolia* | Leguminosae | 1.9 | (Leandro et al., 2016) |
| 7 | *Gymnema sylvestre* | Apocynaceae | 1 | (Dineshbabu et al., 2015) |
| 8 | *Himatanthusdrasticus* | Apocynaceae | 1 | (Figueiredo et al., 2017) |
| 9 | *Leucas aspera* | Lamiaceae | 1.7 | (Dineshbabu et al., 2015) |
| 10 | *Mikania glomerata* | Compositae | 1.5 | (Carrara et al., 2017) |
| 11 | *Pogostemon heyneanus* | Lamiaceae | 1.7 | (Rubini et al., 2018) |
| 12 | *Scutellaria baicalensis* | Lamiaceae | 1.7 | (Leung et al., 2016) |
| 13 | *Vitex negundo* | Lamiaceae | 1.7 | (Dineshbabu et al., 2015) |
| 14 | *Withania somnifera* | Solanaceae | 1.4 | (Malaikozhundan et al., 2020) |
| 15 | Mangrove-derived endophytic fungus<br>*Eurotium chevalieri* KUFA 0006 | Fungi | NA | (Zin et al., 2017) |
| Mixture of extracts |  |  |  |  |
| 16 | *Carotae fructus*, *Arecae semen*, *Granati pericarpium*,<br>*Omphalia lapidescens*, *Coptidis rhizoma*, *Cyrtomii*<br>*rhizoma*, *Meliae cortex*, *Platycladi cacumen*, *Portulacae*<br>*herba*, *Andrographitis herba*, *Radix aucklandiae* | Several families | NA | (Campbell & Gilbert, 2019) |

The presumptive drug target of MRSA biofilm is dehydroxysqualene synthase which produces staphyloxanthin, its main biofilm component [162, 163, 164, 53, 115]. Essential oils are also used to treat infections against gram negative bacteria relevant in veterinary in the Syn-NPs-CAAs format [5]. Limonene and other essential oils, as in Table 1, may be obtained by green extraction methods, they evaporate with time, and they can be detected by electronic noses, which can facilitate quality control in the food industry. Other type of components could be also included in antimicrobial formulations or in the design of plastic polymers that allow for virulence factors modulation [15, 165, 89, 121, 166, 148].

### Increasing Bioavailability and Stimulation of the Host’s Immune System

Several studies using host-pathogen models reported that the enhancement of the antibiotic activity was related to the host’s metabolism, *e*.*g*., stimulation of the immune system or increasing bioavailability. Beyond whole one type cells assays, rodent animal models and mammalian cultured cells allows for detection of such interactions [29, 30, 31, 32, 167, 33, 168, 169, 94, 122, 170].

Those findings highlights the importance of this type of experiments to detect the induction of favorable host-pathogen interactions, at same time highlights the need to access and to develop high-through-put protocols that do not necessarily involves animals but that still can be a probe for those interactions.

### Techniques, Methods and Approaches

Several approaches and techniques are being developed in order to find antibiotic enhancers. Efforts toward rationale design drug discovery are ongoing, such as natural products inspired fragment based approach [171] and SARS studies [172]. And non-targeted mass spectrometry analyses [173] and other metabolomic based methods (biochemometric) [42] are among several platforms for high-through-put bioassays are being proposed [174, 175, 176, 161, 177].

If plant extracts are going to be used as the commercial formulation, their quality control is a key stone for its success. Dettweiler et al., proposed the Extract Fractional Inhibitory Index—EFICI as quality control method of the extract, testing the actual bioactivity of the extract instead of its main components [178].

### Raw materials

Examples of research lines related to circular economy are the synergistic effects of CADS with olive leaf phenols [127], essential oil of melon peel [99], anthocyanins from wine by-products [179], and with metabolites from tobacco waste [180], fruit waste material [99].

Nutrient additives for animals food and soil fertilizers based on plant extracts are attracting interest, they not necessarily contain CADs, but their application eventually leads to reducing the amount of CADs utilized. In the case of animal feed additives. They stimulate the immune system thus potentially reducing the impact of infections and amount of CADs applied. For the case of fertilizers, they not only enrich the soil’s nutrient composition but they can improve the soil’s microbiota for further crop planting [94, 181, 106, 182, 183, 184, 185].

Another approach is the derivatization of natural products by linking them with other privileged scaffolds to improve their potency and ADME properties. For instance, azole derivatives carvacrol and naphtoquinones have been effective *in vitro* against gram positive and negative strains [146] and curcumin derivatives have been prepared aiming to improve their bioavailability [186].

## Conclusions and Final Remarks

Future work would involve to study more natural products and to develop materials that contain antibiotic enhancers. For example: **a)** to investigate more species and to characterize their extracts; **b)** to expand the purified natural products by either testing more NPs or derivatizing those already identified as drug hits, e.g., for absorption improvement; **c)** to systematically explore of plant genus or families; **d)** to develop more test to explore inhibition of virulent factors; **e)** to prepare polymers and films with bioactive natural products for in-field test of their effectivity; **f)** to define formulations to inhibit bacterial growth, that include antibiotic enhancers such as flavonoids, polyphenols or essential oils.

The mixtures of natural products and commercially available antibiotics already shown synergistic effects against opportunistic bacteria relevant in health care, food and plant industries, and several patented formulations related to toothpastes and beauty products are starting to emerge. Those trends grant further research, development and innovation in the food industry and health care systems.

Further activity in this area can led to the surge of non-traditional circular economies around certain species and even to consolidate as an additional tool to manage the resistant bacteria era.

## Data Availability

All data produced in the present study are available upon reasonable request to the authors

## Data Availability

All data produced in the present study are available upon reasonable request to the authors

## Acknowledgements

Authors thanks to University at Buffalo library. LN-V thanks to Chemistry Department Faculty, especially Dr. Troy Wood,; to Dr. Anthony Campagnari and Dr. Nicole Luke from Microbiology and Immunolgy Department, from University at Buffalo, New York,; and to Arturo Solis for sharing their expertise.

## Abbreviations

Text for this section.

## Availability of data and materials

Excel base document and tables are availabe upon request.

## Ethics approval and consent to participate

This study did not involved human subjects information or experiments with animal models.

## Consent for publication

All the authors consent for publication in the Future Journal of Pharmaceutical Sciences.

## Authors’ contributions

LN-V performed the literature search, statistic analysis, and prepared manuscript; SBB extracted and verified the botanical data; RAMS elaborated several tables; RC prepared several structures.

## Additional Files

Excel file with raw data is available upon request.

